# Characterizing the countrywide epidemic spread of influenza A(H1N1)pdm09 virus in Kenya between 2009 and 2018

**DOI:** 10.1101/2021.03.30.21254587

**Authors:** D. Collins Owuor, Zaydah R. de Laurent, Gilbert K. Kikwai, Lillian M. Mayieka, Melvin Ochieng, Nicola F. Müller, Nancy A. Otieno, Gideon O. Emukule, Elizabeth N. Hunsperger, Rebecca Garten, John R. Barnes, Sandra S. Chaves, D. James Nokes, Charles N. Agoti

## Abstract

**Background:** The spatiotemporal patterns of spread of influenza A(H1N1)pdm09 viruses on a countrywide scale are unclear in many tropical/subtropical regions mainly because spatiotemporally representative sequence data is lacking.

**Methods:** We isolated, sequenced, and analyzed 383 influenza A(H1N1)pdm09 viral genomes isolated from hospitalized patients between 2009 and 2018 from seven locations across Kenya. Using these genomes and contemporaneously sampled global sequences, we characterized the spread of the virus in Kenya over several seasons using phylodynamic methods.

**Results:** The transmission dynamics of influenza A(H1N1)pdm09 virus in Kenya was characterized by: (i) multiple virus introductions into Kenya over the study period, although these were remarkably few, with only a few of those introductions instigating seasonal epidemics that then established local transmission clusters; (ii) persistence of transmission clusters over several epidemic seasons across the country; (iii) seasonal fluctuations in effective reproduction number (*R*_e_) associated with lower number of infections and seasonal fluctuations in relative genetic diversity after an initial rapid increase during the early pandemic phase, which broadly corresponded to epidemic peaks in the northern and southern hemispheres; (iv) high virus genetic diversity with greater frequency of seasonal fluctuations in 2009-11 and 2018 and low virus genetic diversity with relatively weaker seasonal fluctuations in 2012-17; and (v) virus migration from multiple geographical regions to multiple geographical destinations in Kenya.

**Conclusion:** Considerable influenza virus diversity circulates within Africa, as demonstrated in this report, including virus lineages that are unique to the region, which may be capable of dissemination to other continents through a globally migrating virus population. Further knowledge of the viral lineages that circulate within understudied low-to-middle income tropical and subtropical regions is required to understand the full diversity and global ecology of influenza viruses in humans and to inform vaccination strategies within these regions.

## INTRODUCTION

The novel influenza A(H1N1)pdm09 virus strain emerged in North America during March-April 2009, spread rapidly among humans, and developed into the first human pandemic of the 21^st^ century [1-4]. By July 2009, 168 countries reported a total of 162,300 laboratory-confirmed cases and over 1,100 human deaths [5-7]. Subsequently, it was estimated that over 123,000 deaths globally from March to December 2009 were associated with A(H1N1)pdm09 virus infection [8]. The A(H1N1)pdm09 virus displaced seasonal A(H1N1) virus and has continued to circulate as a seasonal virus in subsequent years, causing annual seasonal epidemics alongside influenza A(H3N2) and B viruses globally [9-12], including Kenya [13-17].

The global surveillance of influenza viruses has resulted in the generation of an extensive collection of geographically and temporally comprehensive virus sequence data, which has provided an opportunity to explore the drivers of global spread of influenza viruses [18-22]. For instance, genomic analysis found that multiple independent introductions of genetically distinct A(H1N1)pdm09 virus lineages occurred in most countries, for example, in the United Kingdom (UK) [9, 23]. However, only two of the many lineages that were introduced at the start of the pandemic in the UK were detected there six months later [9, 23]. Elsewhere, a pair of A(H1N1)pdm09 virus transmission chains appear to have persisted in west Africa for almost two years [24]. After the A(H1N1)pdm09 virus became endemic, such spatiotemporal patterns of spread of A(H1N1)pdm09 viruses on a countrywide scale as reported for the UK and west Africa have, however, not been determined in many tropical/subtropical regions where virus circulation patterns do not show clear seasonality as is observed in temperate countries [25, 26]. This is often due to insufficient spatiotemporally representative sequence data [27, 28]. A more complete understanding of the regional, as well as the global spread of influenza viruses, however, requires deeper and wider sampling from understudied tropical/subtropical regions [28].

In Kenya, influenza surveillance activities reported at least four separate introductions of A(H1N1)pdm09 virus into the country during the pandemic in 2009, with the first laboratory-confirmed case reported on June 29, 2009 [29, 30]. In an effort to fill the gap in studies that seek to describe the transmission of influenza viruses in tropical settings, we studied the Kenya-wide patterns of introduction and spread of A(H1N1)pdm09 virus since it was introduced into the local population in 2009. We isolated, sequenced, and analyzed 383 A(H1N1)pdm09 virus codon-complete genome sequences sampled between 2009 and 2018 from seven locations in Kenya alongside contemporaneously sampled global sequences, to investigate the introduction and spread of A(H1N1)pdm09 viruses to Kenya.

## MATERIALS AND METHODS

### Sample sources and molecular screening

Samples analyzed in this study were collected from two health facility-based surveillance networks. The first involved continuous countrywide surveillance for influenza through severe acute respiratory illness (SARI) sentinel hospital reporting. This was undertaken at six sites supported by the United States Centers for Disease Control and Prevention (CDC): Kenyatta National Hospital (KNH), Nakuru County and Referral Hospital (CRH), Nyeri CRH, Kakamega CRH, Siaya CRH, and Coast General Teaching and Referral Hospital (**S1 Figure**) [13, 14, 16, 31]. The second was the pediatric viral pneumonia surveillance undertaken at the Kilifi County Hospital (KCH) (**S1 Figure**) [32].

In the CDC-supported surveillance sites (CDC-Kenya), a total of 41,685 nasopharyngeal/oropharyngeal (NP/OP) swab samples were collected from inpatient admissions of all ages from June 2009 through December 2018. Samples were stored in viral transport medium (VTM) at −80°C prior to processing [13]. Of these, 41,102 swabs were tested for influenza A virus (IAV), with positive samples subsequently subtyped for influenza A(H1N1)pdm09 and A(H3N2) viruses using real-time reverse transcription polymerase chain reaction (RT-PCR) [13, 14, 16]. A total of 1,307 (3.2%) A(H1N1)pdm09 virus positive samples were obtained. Of these, 418 (31.9%) A(H1N1)pdm09 virus positive samples were selected for inclusion in this analysis based on: (a) RT-PCR cycle threshold (Ct) of <35.0 (as proxy for high viral load); (b) adequate sample volume for RNA extraction (>140 μL); and (c) balanced distribution of samples across the different surveillance sites and years.

In the second facility-based surveillance undertaken at KCH from January 2009 through December 2018, a total of 6,147 NP/OP swab samples were collected and tested from children <5 years admitted with syndromic severe or very severe pneumonia [15, 32]. Samples were stored in VTM at −80°C prior to molecular screening. Samples were screened for a range of respiratory viruses, including IAV, using a multiplex reverse transcription (RT)-PCR assay employing Qiagen QuantiFast RT-PCR kit (Qiagen) [33]. A (RT)-PCR Ct of <35.0 was used to define virus-positive samples [33]. A total of 157 (2.6%) IAV virus positive specimens were identified for inclusion from KCH; however, these were not subsequently subtyped for A(H1N1)pdm09 and A(H3N2) viruses [32]. Therefore, all were utilized in the current analysis.

### RNA extraction and multi-segment real-time PCR (M-RTPCR) for IAV

Viral RNA extraction and M-RTPCR was conducted as previously described [34]. Briefly, viral nucleic acid extraction from IAV and A(H1N1)pdm09 virus positive samples (Ct <35.0) was performed using the QIAamp Viral RNA Mini Kit (Qiagen). Ribonucleic acid (RNA) was reverse transcribed, and the complete coding region of IAV genome was amplified in a single M-RTPCR using the Uni/Inf primer set [35] in 25 μL PCR reactions. Successful amplification was evaluated by running the products on 2% agarose gel and visualized on a UV transilluminator after staining with RedSafe Nucleic Acid Staining solution (iNtRON Biotechnology Inc.,).

### IAV next-generation sequencing (NGS) and virus genome assembly

Following PCR, amplicons were purified, quantitated and normalized as previously described [34]. Briefly, the amplicons were purified with 1X AMPure XP beads (Beckman Coulter Inc.,), quantified with Quant-iT dsDNA High Sensitivity Assay (Invitrogen), and normalized to 0.2 ng/μL. Indexed paired end libraries were generated from 2.5 μL of 0.2 ng/μL amplicon pool using Nextera XT Sample Preparation Kit (Illumina) following the manufacturer’s protocol. Amplified libraries were purified using 0.8X AMPure XP beads, quantitated using Quant-iT dsDNA High Sensitivity Assay (Invitrogen), and evaluated for fragment size in the Agilent 2100 BioAnalyzer System using the Agilent High Sensitivity DNA Kit (Agilent Technologies). Libraries were then diluted to 2nM in preparation for pooling and denaturation for running on the Illumina MiSeq (Illumina). Pooled libraries were sodium hydroxide denatured, diluted to 12.5 pM and sequenced on the Illumina MiSeq using 2 × 250 bp paired end reads with the MiSeq v2 500 cycle kit (Illumina). Five percent Phi-X (Illumina) spike-in was added to the libraries to increase library diversity by creating a more diverse set of library clusters. Contiguous (contigs) nucleotide sequence assembly from the sequence data was carried out using the FLU module of the Iterative Refinement Meta-Assembler (IRMA) [36] using IRMA default settings: median read Q-score filter of 30; minimum read length of 125; frequency threshold for insertion and deletion refinement of 0.25 and 0.6, respectively; Smith-Waterman mismatch penalty of 5; and gap opening penalty of 10. All generated sequence data were deposited in the Global Initiative on Sharing All Influenza Data (GISAID) EpiFlu™ database (https://platform.gisaid.org/epi3/cfrontend) using accession numbers listed in the metadata file in the report’s GitHub repository https://github.com/DCollinsOwuor/H1N1pdm09_Kenya_Phylodynamics/tree/main/Data/.

### Collation of contemporaneous global sequence dataset

Global comparison datasets for influenza A(H1N1)pdm09 virus were retrieved from the GISAID EpiFlu™ database (https://platform.gisaid.org/epi3/cfrontend). The datasets were prepared to determine the relatedness of the viruses in this report to those circulating around the world thus understand their global context. Only sequences with complete coding sequences sampled between March 2009 and December 2018 were included to improve the phylogenetic resolution of the analyses. The data were organized into a Microsoft Excel database which also stored the associated metadata (country of origin, date of isolation, subtype, and sequence length per segment). In-house python scripts were used in the extraction and manipulation of the data. Additionally, sequences were binned by calendar year for temporal analysis. A final dataset of 1,587 global sequences sampled between April 2009 and December 2018 was available (numbers in parenthesis indicate number of sequences): Africa (155); Asia (372); Europe (326); North America (356); South America (181); and Oceania (197). The accession numbers for the global dataset are available in the GISAID acknowledgement table in the report’s GitHub repository, https://github.com/DCollinsOwuor/H1N1pdm09_Kenya_Phylodynamics/tree/main/Data/.

### Phylogenetic analysis

Consensus nucleotide sequences were aligned and translated in AliView v1.26 [37] and the individual gene segments of Kenyan sequences were concatenated into codon-complete genomes using SequenceMatrix [38]. Phylogenetic trees of A(H1N1)pdm09 virus genomes from Kenya and contemporaneously sampled global sequences were reconstructed with maximum-likelihood and bootstrap analysis of 1,000 replicates. The best-fit nucleotide substitution models were inferred using IQ-TREE v1.6.11 [39, 40] and those chosen by the Bayesian Information Criterion for each concatenated virus genome implemented. The phylogenetic trees were visualized and annotated using Figtree v1.4.4 (http://tree.bio.ed.ac.uk/software/figtree/) and ggtree [41]. The full-length hemagglutinin (HA) codon sequences of all viruses were used to characterize A(H1N1)pdm09 virus strains into genetic groups (i.e., clades, subclades, and subgroups) based on the European CDC Guidelines (https://www.ecdc.europa.eu/en/seasonal-influenza/surveillance-and-disease-data/influenza-virus-characterisation) and Phylogenetic Clustering using Linear Integer Programming (PhyCLIP) [42]. Representative reference sequences for genetic clade assignment were included.

### Estimating population dynamics of A(H1N1)pdm09 virus in Kenya from local transmission clusters

To analyze the introduction and local spread of A(H1N1)pdm09 virus in Kenya, different local transmission clusters, defined as groups of sequences that originate from a single introduction into Kenya, were first identified. Ancestral state reconstruction of internal nodes based on Kenyan and global sequences was used to infer these transmission clusters using a maximum-likelihood approach in TreeTime [43]. To do so, all the sequences were assigned into two discrete location states: (i) Kenyan - all Kenyan sequences and (ii) non-Kenyan – all sequences from anywhere else from the globe. All sequences that cluster together are considered to be in the same transmission cluster if their common ancestral nodes are inferred to be in Kenya whereas each individual transmission cluster is a result of a single introduction into Kenya. Clusters were named to reflect their placement within global genetic clades 1-7, for example, KENI-GC7 indicates Kenya cluster I viruses within global genetic clade 7. These clusters were then used to analyze the spread of A(H1N1)pdm09 virus in Kenya using two phylodynamic approaches. First, the effective reproduction number (*R*_e_), which is the average number of secondary cases generated by an infection, was estimated using a birth-death skyline (BDSKY) analysis [44], where all individual transmission clusters are assumed to be independent observations of the same process with the same parameters [45]. Second, the relative genetic diversity (effective virus population size over time) was estimated using Gaussian Markov random field (GMRF) coalescent smoothing of the effective population size [46].

### Spatial dynamics of A(H1N1)pdm09 virus in Kenya

We conducted phylogeographic analysis to assess virus spread among three geographical regions of Kenya: (i) central Kenya - Nairobi, Nakuru, Nyeri; (ii) western Kenya – Siaya and Kakamega; (iii) and coastal Kenya - Mombasa, and Kilifi (**S1 Figure**) using methods implemented in BEAST v1.10.4 package [47]. The analysis was implemented with an asymmetrical discrete trait approach and applied the Bayesian stochastic search variable selection (BSSVS) model [48]. Phylogeographic inferences were visualized with the Spatial Phylogenetic Reconstruction of Evolutionary Dynamics using Data-driven Documents (SPREAD3) software v0.9.7.1c [49]. To visualize the geographic spread of the virus over time, a D3 file was generated using SPREAD3 v0.9.7.1c. A Kenya geo.json file was used for visualization and resulting log files were used to calculate Bayes factor (BF) values for significant dispersal rates between discrete locations.

## RESULTS

### IAV sequencing and genome assembly

Among the 418 A(H1N1)pdm09 virus positive samples that were available from CDC-Kenya, 414 (99.1%) passed the pre-sequencing quality control checks (see **IAV next-generation sequencing and virus genome assembly** above) and were loaded onto the MiSeq, **S2 Figure**. This yielded 344 (83.1%) codon-complete and 66 (15.9%) partial A(H1N1)pdm09 virus genomes. For this report, only the 344 codon-complete A(H1N1)pdm09 virus genome sequences were used. Among the 157 IAV positive specimens available from KCH, 94 (59.9%) that passed pre-sequencing quality control checks were loaded onto the MiSeq with corresponding success in generating 45 (47.9%) A(H1N1)pdm09 virus (39 codon-complete and 6 partial) and 49 (52.1%) A(H3N2) virus genomes (46 codon-complete and 3 partial). For this report, only the 39 codon-complete A(H1N1)pdm09 virus sequences were used, **S2 Figure**.

### Spatiotemporal distribution of sequenced samples

The A(H1N1)pdm09 virus was detected throughout the study period in Kenya, with the number of observed cases fluctuating from year-to-year, **Figure 1, Panel A**. Different locations experienced epidemic peaks in different years. However, there were A(H1N1)pdm09 virus detections in all sites, except in mid-2010, 2012-2013, and 2016 when there was little transmission of A(H1N1)pdm09 virus throughout the different sites, **Figure 1, Panel A**. The proportion of sequenced samples roughly reflected the overall distribution of positives that were detected from each site, **Figure 1, Panel A**. All A(H1N1)pdm09 virus genetic groups were detected in most sites, with their majority detected in 5 or 6 of the 7 sites, which suggests that these lineages were in circulation in most of the sites in Kenya without geographical restrictions during the study period, **Figure 1, Panel B**. Phylogenetic analysis showed that the sequenced 383 A(H1N1)pdm09 viruses from Kenya comprised 7 genetic groups: clade 7 (n=97, 25.3%), clade 6 (n=132, 34.5%), subclade 6C (n=10, 2.6%), subclade 6B (n=47, 12.3%), subclade 6B.1 (n=38, 9.9%), subgroup 6B.1A (n=57, 14.9%), and subgroup 6B.1A1 (n=2, 0.5%), **Figure 2**. These detections varied by surveillance year: clade 7 – 2009-12; clade 6 – 2009-11; subclade 6C – 2013-14; subclade 6B – 2014-16; subgroup 6B.1 – 2015-16; subgroup 6B.1A and subgroup 6B.1A1 – 2018 (**Figure 1, Panel C**).

**Figure 1:**
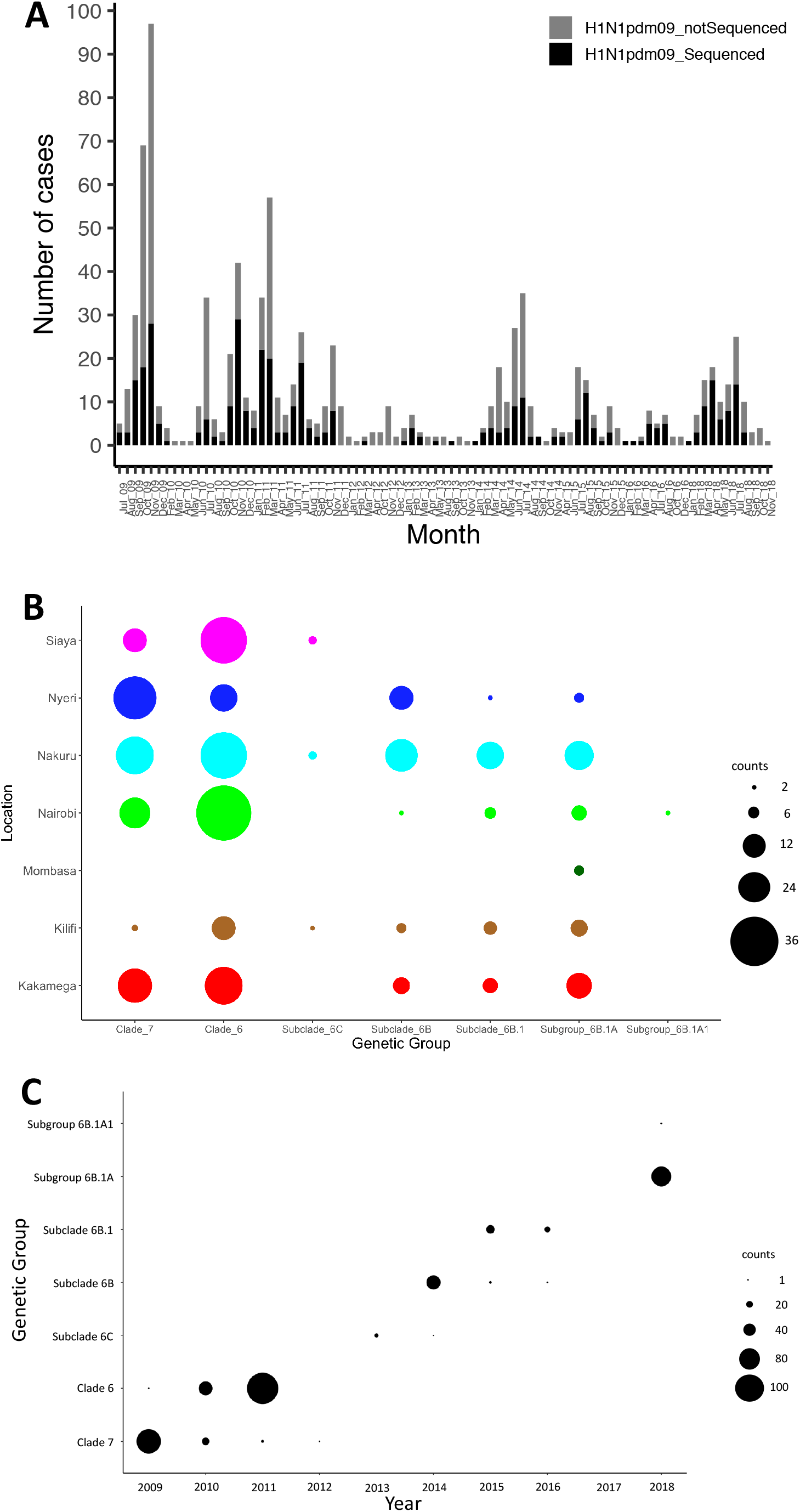
Distribution of influenza A(H1N1)pdm09 virus in Kenya, 2009-2018. A Bar plot showing number of A(H1N1)pdm09 virus positive samples and sequenced positive samples by month between July 2009 and November 2018 in Kenya. All collected A(H1N1)pdm09 virus positive samples and sequenced samples are indicated by color (all positive samples in gray – H1N1pdm09_notSequenced; sequenced samples in black - H1N1pdm09_Sequenced) as shown in the color key. **B** Bubble plot showing the distribution of genetic groups by location in Kenya, 2009 – 2018. **C** Distribution of genetic groups by surveillance year in Kenya, 2009 - 2018. The size of the circle in panels **B** and **C** is proportional to number of samples as shown in the counts key for the figures.

**Figure 2:**
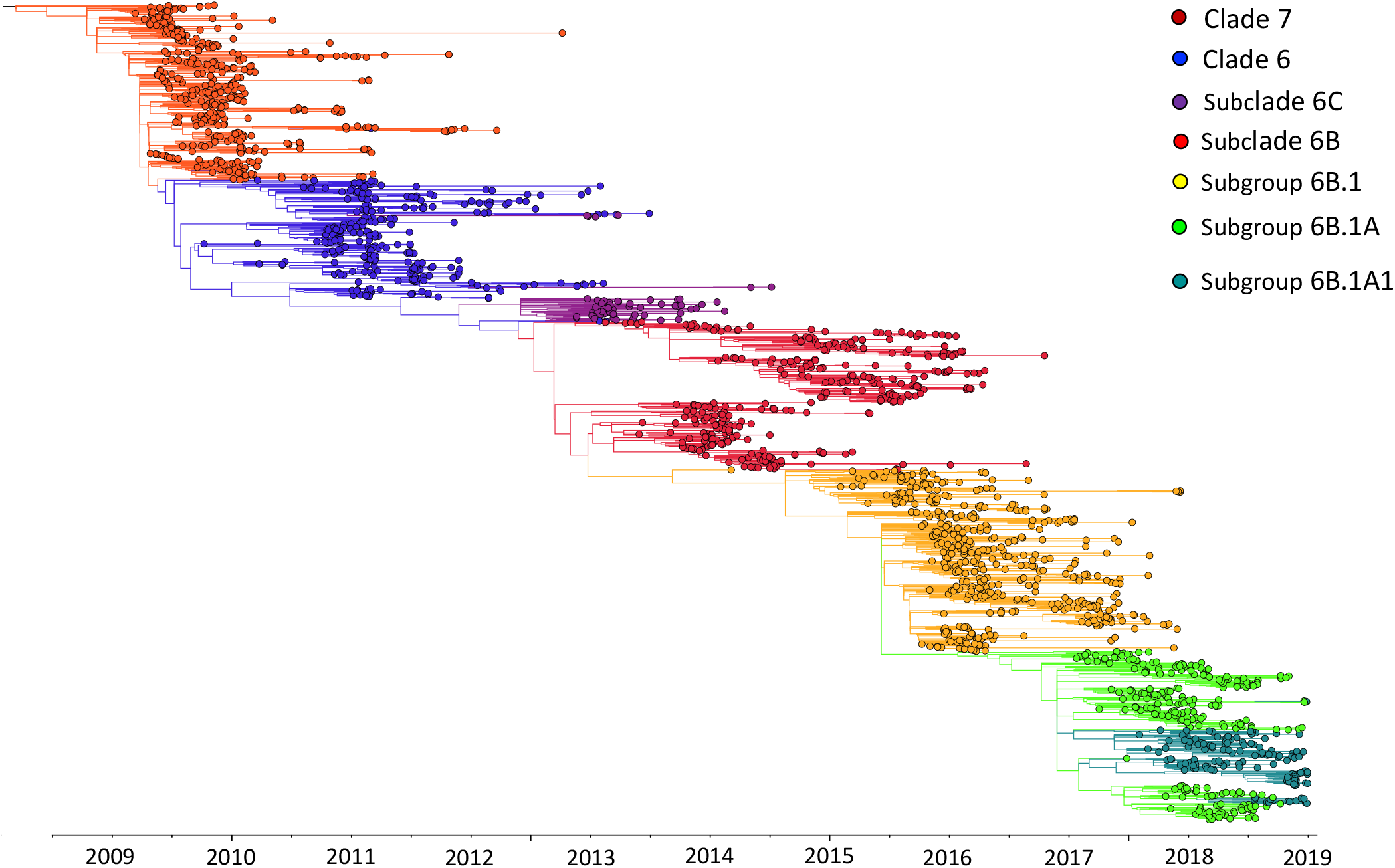
Maximum-likelihood phylogenetic tree of 1,970 influenza A(H1N1)pdm09 virus sequences from Kenya and contemporaneously sampled global locations collected between 2009 and 2018. This is a time-calibrated phylogeny with time shown on the x-axis. Branches are colored based on genetic group membership as shown in the color key.

### Patterns of introduction of A(H1N1)pdm09 viruses into Kenya and local transmission clusters

We first assessed how the 383 sequences from Kenya compare to 1,587 sequences sampled from around the world between 2009 and 2018 by inferring their phylogenies. The Kenyan genomes span the existing global diversity (**Figure 3**), which suggests exchange (most likely introductions into Kenya) of viruses with other areas around the globe. For example, phylogenetic tree trunk viruses predominantly originated from North America in 2009, consistent with the origins of A(H1N1)pdm09 virus in North America in 2009, which seeded global viruses in 2009-10. Subsequently, Asia and Europe appeared to be the major source population in 2010-13 and 2014-18, respectively, **Figure 3**. The three geographical regions also represent sources of introduction of A(H1N1)pdm09 virus into Kenya in 2009, 2010-13 and 2014-18. Based on reconstruction of geographic ancestry, Kenyan sequences grouped into local transmission clusters within the global diversity, **Figure 3**. We inferred 30 transmission clusters (KENI-GC7 to KENXXX-GC6B.1A), **Figure 3**, which suggests that the sampled sequences were the result of 30 independent introductions from areas outside of Kenya. However, despite the relatively strong clustering of Kenyan sequences into transmission clusters, there were relatively fewer introductions over the study period since for most years, only a few introductions caused epidemics. To investigate if this number is a strict lower bound for the introductions, we used random subsets of the 383 A(H1N1)pdm09 virus sequences from Kenya to re-estimate the number of introductions, **Figure 4**. We find that the number of estimated introductions starts to flatten slightly with the number of sequences subsampled but is still growing. This saturating relationship suggests that the study is not grossly underestimating the full number of A(H1N1)pdm09 virus variants in this report. Seven of the thirty transmission clusters appeared to have persisted over several epidemic seasons (**Table 1**), which provides evidence for multi-year persistence of individual A(H1N1)pdm09 virus transmission clusters in a specific locality. All the seven transmission clusters that persisted between 2009 and 2018 consisted of viruses sampled across Kenya, which suggests that A(H1N1)pdm09 virus persistence in the country was not constrained geographically.

**Figure 3:**
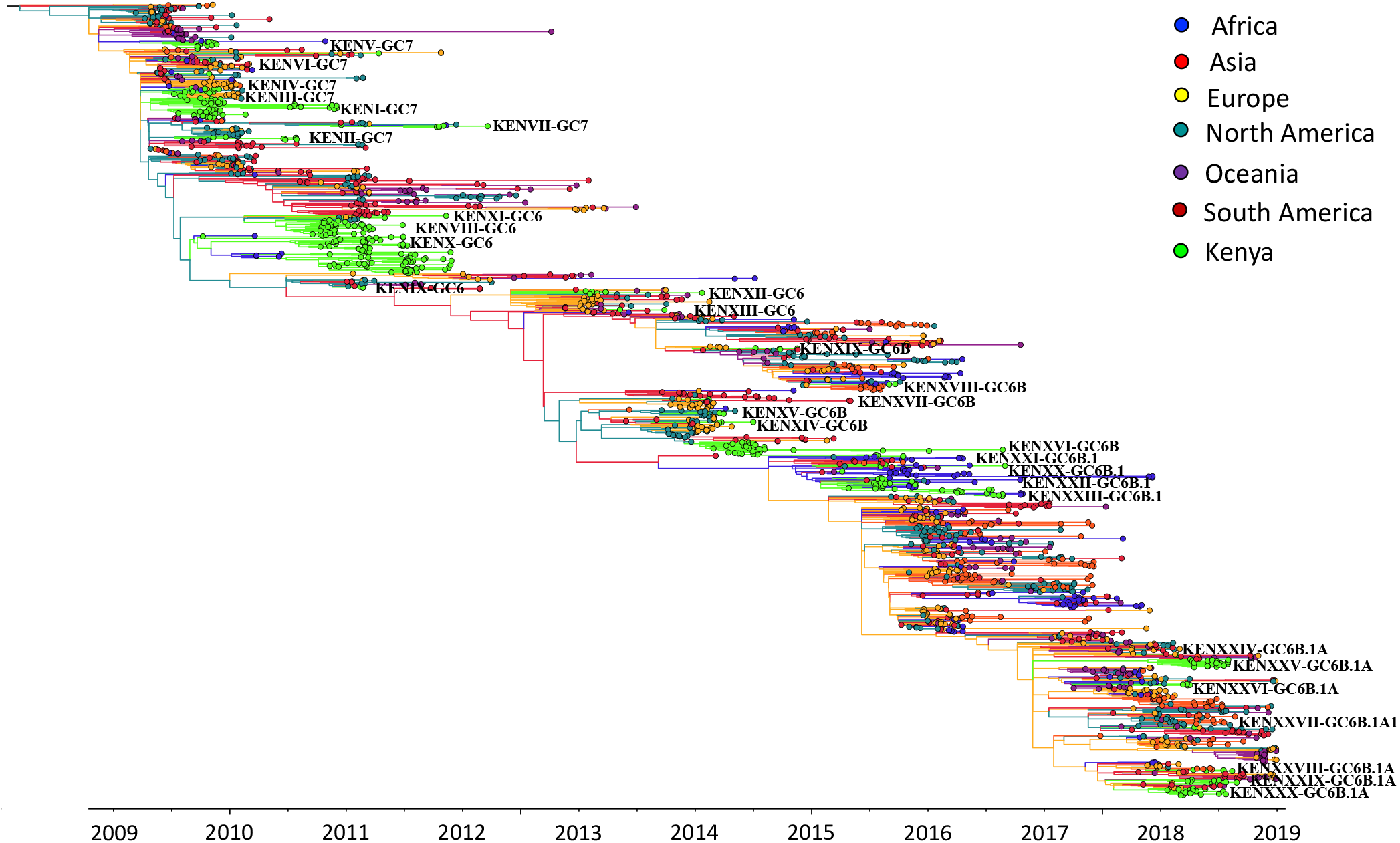
Time-resolved maximum-likelihood phylogenetic tree of Kenyan and contemporaneously sampled global sequences collected between 2009 and 2018 showing continent of sequence sampling and Kenyan transmission clusters. Unique Kenyan clusters are labeled with the prefix KEN, followed by cluster grouping and genetic group, for example, KENI-GC7 indicates Kenyan cluster I viruses, which fall within global genetic clade 7. The branches are colored based on continent of sampling as shown in the color key. Additionally, the trunk locations are inferred and colored by continent, which is based on geographic ancestry analyses of sampled sequences to indicate influenza A(H1N1)pdm09 virus origins.

**Figure 4:**
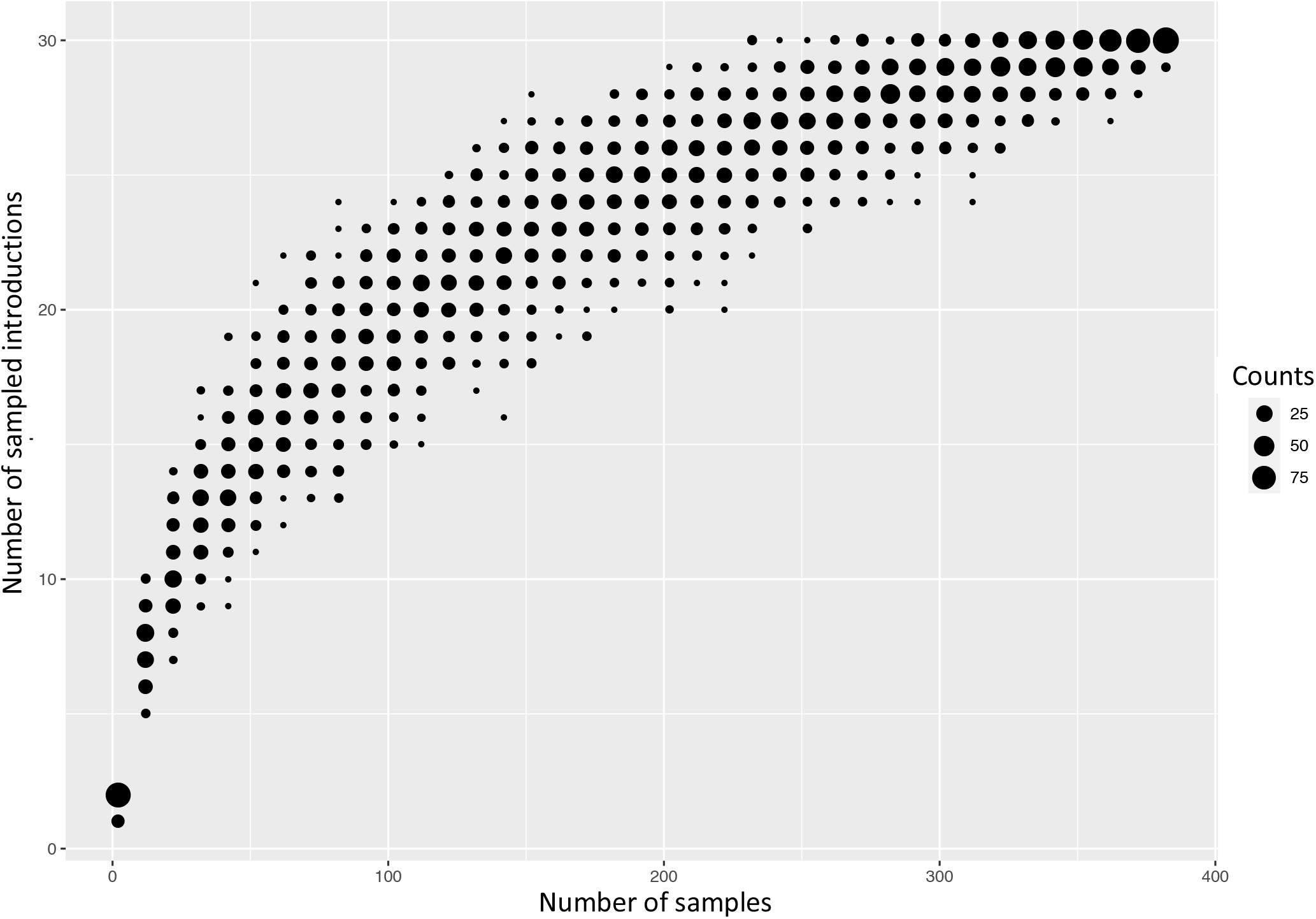
Number of introductions of A(H1N1)pdm09 viruses into Kenya depending on how many random sequences from Kenya are used to infer introductions.

**Table 1:** Patterns of persistence of A(H1N1)pdm09 virus transmission clusters in Kenya.

| †Cluster | Clade | Sequences | From | Circulation | Duration of spread | Locations detected |
| --- | --- | --- | --- | --- | --- | --- |
| KENI-GC-7 | 7 | 49 | Europe | July 2009 to December 2010 | 1.5 years | Nairobi; Nakuru; Nyeri; Siaya; Kilifi |
| KENVII-GC7 | 7 | 6 | North America | December 2010 to March 2012 | 1.3 years | Nairobi; Nakuru; Kakamega |
| KENVIII-GC6 | 6 | 39 | North America | October 2010 to June 2011 | 9 months | Nairobi; Nakuru; Kakamega; Nyeri; Siaya; Kilifi |
| KENX-GC6 | 6 | 88 | North America | October 2009 to November 2011 | 2.2 years | Nairobi; Nakuru; Kakamega; Nyeri; Siaya; Kilifi |
| KENXII-GC6C | 6C | 8 | Europe | January 2013 to January 2014 | 1 year | Nakuru; Siaya |
| KENXVI-GC6B | 6B | 34 | North America | February 2014 to August 2016 | 2.5 years | Nairobi; Nakuru; Kakamega; Nyeri; Kilifi |
| KENXXIII-GC6B.1 | 6B.1 | 34 | Europe | April 2015 to August 2016 | 1.3 years | Nairobi; Nakuru; Kakamega; Nyeri; Kilifi |

† Cluster, name of transmission cluster; Clade, genetic group membership; Sequences, number of sequences in cluster; From, inferred geographical source of virus introduction; Circulation, duration of circulation of the cluster in Kenya; Locations detected, Kenyan locations where clusters were detected.

### Estimating population dynamics of A(H1N1)pdm09 virus in Kenya from the local transmission clusters

In order to quantify the amount of local transmission based on the local transmission clusters, we estimated the effective reproduction number to be between 1 and 1.5 throughout the study period (**Figure 5, Panel A**). We inferred seasonal fluctuations in *R*_e_ between 2009 and 2018, with annual peaks in *R*_e_ usually occurring at the end of the year and annual drops in *R*_e_ following the annual peaks, with the estimated median often being below 1, **Figure 5, Panel A**. The low *R*_e_ values throughout 2009-18 are consistent with the occurrence of smaller epidemics in Kenya throughout the study period. Coalescent reconstruction of A(H1N1)pdm09 virus occurrence in Kenya revealed: (i) seasonal fluctuations in relative genetic diversity after an initial rapid increase during the early pandemic phase, which broadly corresponded to epidemic peaks in the northern and southern hemispheres; (ii) higher genetic diversity (genetic diversity >10), with greater frequency of seasonal fluctuations observed during 2009-2011 and 2018; and (iii) lower genetic diversity (variation occurring between 1 and 10 in genetic diversity), with relatively weaker seasonal fluctuations sustained from 2012-2017, **Figure 5, Panel B**.

**Figure 5:**
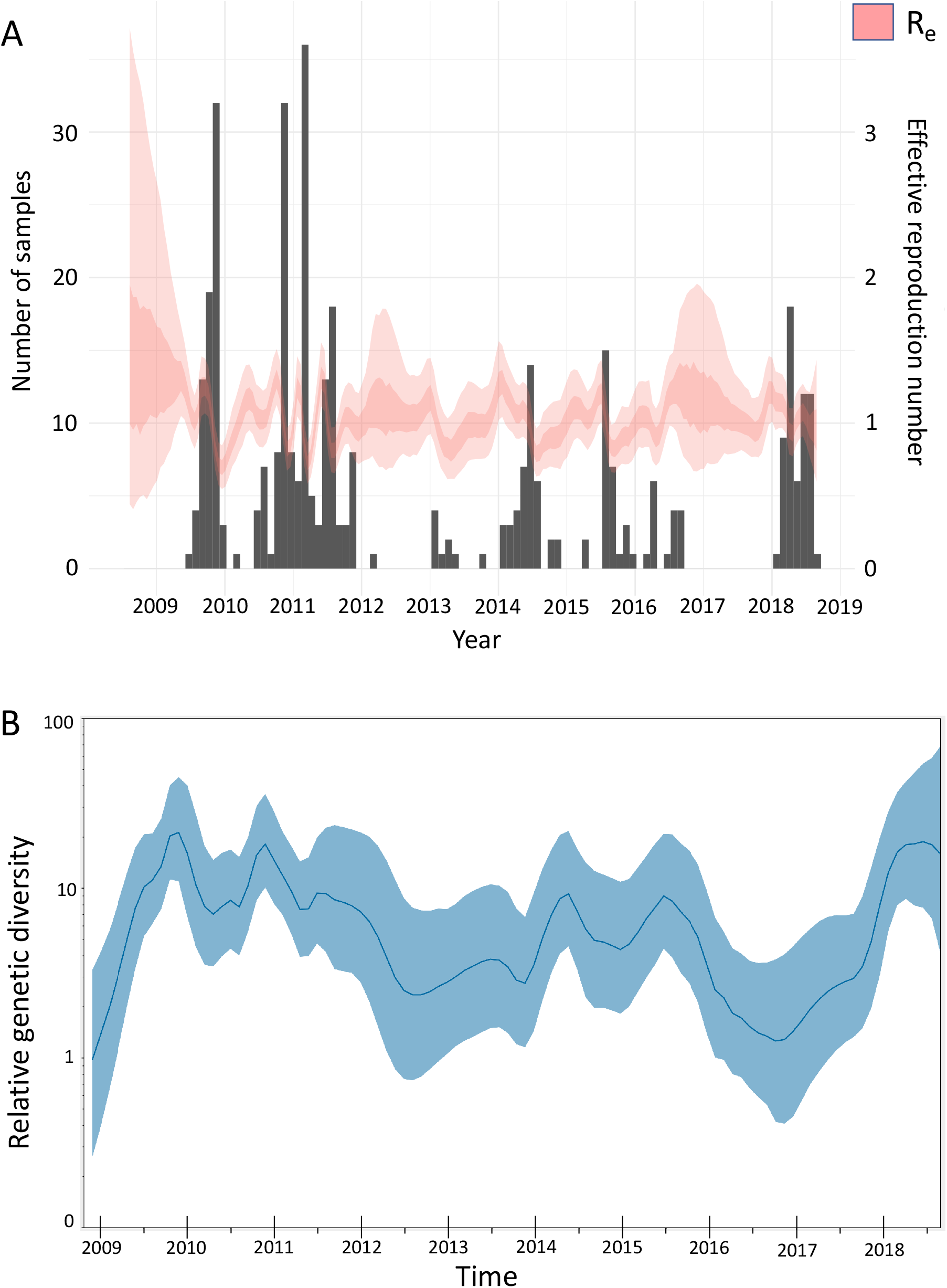
Population dynamics of influenza A(H1N1)pdm09 virus from Kenya, 2009-2018. **A** Estimates of the effective reproduction number through time inferred from all local clusters jointly by using BDSKY analysis. The primary y-axis shows the number of sequenced samples while the secondary y-axis shows the effective reproduction number (*R*_e_). The dark pink section of the *R*_e_ values is the mean *R*_e_ estimate whereas the light pink margins denote the 95% confidence interval; time in years is shown on the x-axis. **B** Estimates of the relative genetic diversity through time for influenza A(H1N1)pdm09 virus from Kenya, 2009-2018, resolved using GMRF analysis. The dark blue line is the mean estimate, and the blue margin denotes the 95% interval. The relative genetic diversity is shown on the y-axis while time is shown on the x-axis. BDSKY, birth-death skyline; GMRF, Gaussian Markov random field.

### Regional spread of A(H1N1)pdm09 virus in Kenya

To infer the patterns of spread of A(H1N1)pdm09 virus among three geographical regions of Kenya (central Kenya, western Kenya, and coastal Kenya), we summarized and visualized its geographic migration over time based on significant migration rates between the geographical regions. We observed significant migration rates from western Kenya to central and coastal Kenya. Additionally, we observed supported migration rates from coastal Kenya to western and central Kenya, **Figure 6**.

**Figure 6:**
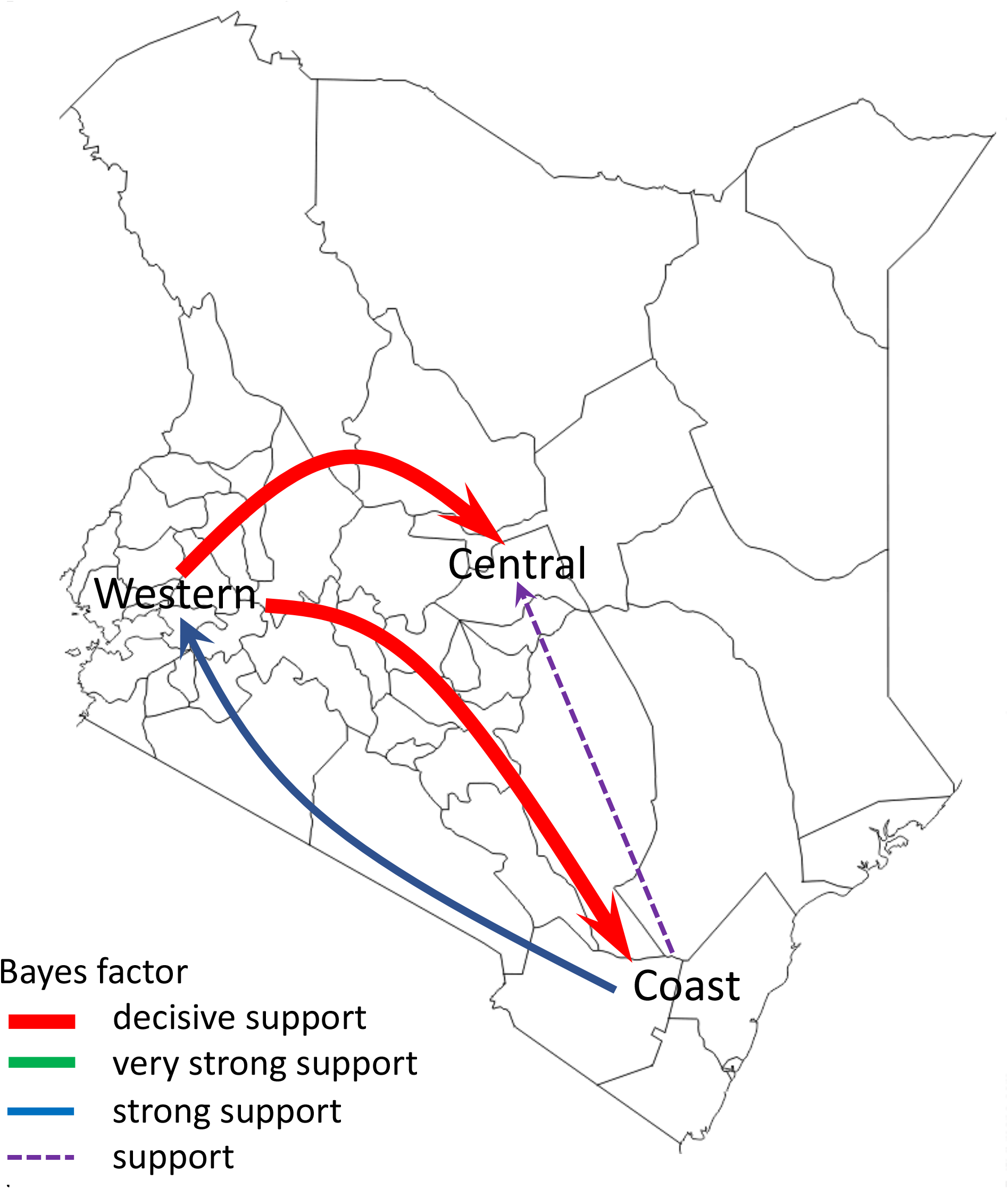
Migration networks of influenza A(H1N1)pdm09 virus reconstructed using sequence data from Kenya, 2009-2018. Asymmetric migration pathways between geographical regions of Kenya (central Kenya, western Kenya, and coastal Kenya) were inferred. Coloured line arrows indicate significant migration routes from one location state to another, while line thickness represents the degree of statistical support. Red arrowed lines are shown to indicate decisive migration routes with Bayes factor (BF) support ≥1000; green lines represent very strongly supported routes with 100≤BF<1000; blue lines indicate strongly supported routes 10≤BF<100; and purple dotted lines indicate supported routes with 3≤BF<10.

## DISCUSSION

We observed multiple A(H1N1)pdm09 virus introductions into Kenya over the study period although these were remarkably few, with only a few of these introductions instigating seasonal epidemics that then established local transmission clusters, some of which persisted over several epidemic seasons across Kenya. Furthermore, we show that the seasonal epidemics were associated with lower number of infections (low estimates of *R*_e_), consistent with estimates from other regions for A(H1N1)pdm09 virus during seasonal epidemics [50], and seasonal fluctuations in virus genetic diversity. Additionally, the spread of A(H1N1)pdm09 virus in Kenya was predominantly driven by virus dispersal from multiple geographical regions to multiple geographical destinations following introduction.

Genomic analysis of virus sequence data from Kenya during the pandemic in 2009 reported the introduction of clade 2 and clade 7 viruses in Kenya, although clade 2 viruses did not circulate beyond the introductory foci while clade 7 viruses disseminated countrywide [51]. Here, through our detailed genomic analysis, we extend these earlier observations to show that clade 7 and clade 6 viruses were introduced into Kenya during the pandemic, disseminated countrywide, and persisted across multiple epidemics in multiple locations as local transmission clusters. A key question in influenza virus evolution and epidemiology is whether viral lineages can persist at low levels of circulation on local and regional scales or whether new virus strains must be continually reseeded from a globally sustained gene pool [24]. The intensive sampling of viruses during the pandemic in 2009-10 enabled the molecular epidemiology of IAV to be examined at such a high resolution that the introduction, persistence and/or fade-out of individual transmission clusters in specific localities could be determined [5, 22]. For example, only two of many United Kingdom-specific A(H1N1)pdm09 virus lineages that persisted between the first and the second pandemic waves of 2009 were detected there six months later [9], while a pair of A(H1N1)pdm09 virus transmission chains appear to have persisted in west Africa for almost 2 years [24]. Our analysis revealed sustained persistence of seven A(H1N1)pdm09 virus transmission clusters for over 2 years, although increased sampling is required to confirm that isolates from other localities are not interspersed within these clusters.

Non-molecular epidemiological studies have hinted at climate-driven patterns of influenza virus spread in Africa, for example, in Kenya [52] and Uganda [53], where climatic factors have been shown to influence the activity of influenza viruses. Therefore, persistence in such African countries might be facilitated by climatic variability, which can generate temporally overlapping epidemics in neighboring regions [24]. Such patterns have been associated with global migration and persistence of influenza viruses in East and Southeast Asia [19]. Our findings support a shifting metapopulation model of circulation of influenza viruses in which viruses may pass through any geographical region for a variable amount of time rather than perpetually circulating in fixed locations, whereby, new virus strains can emerge in any geographical region, with the location of the source population changing regularly [19]. Wider and deeper sampling of viruses from understudied tropical and subtropical regions is therefore required for a more complete understanding of the regional, as well as the global spread of influenza viruses.

Inclusion of regional and global genome sequences deposited in GISAID significantly improved the power of our phylogenomic analyses, which showed that the Kenyan diversity was part of the global continuum. For example, we showed widespread mixing of Kenyan lineages with global viruses from Africa, Asia, Europe, North America, South America, and Oceania. The use of NGS technology to generate virus sequence data from Kenya enables further scrutiny of the available data to answer other key molecular epidemiological questions. For example, the sequencing depth achieved with NGS may allow for analysis of minority variant populations. These might be evolving locally in the region thus undermining the assumption that vaccines matched to globally dominant lineages may necessarily protect against these local lineages.

The study had some limitations. First, the analysis in this report only involved the coding regions of the A(H1N1)pdm09 virus gene segments. Although non-coding regions are considered to be conserved, mutations that affect viral replication may occur and this information may not have been captured in this study. Second, the paucity of sequence data from other African countries limited the analysis of regional patterns of persistence of influenza viruses, since persistence may be facilitated by climatic variability that generates temporally overlapping epidemics in neighboring countries. Lastly, the prioritized samples were selected on the basis of anticipated probability of successful sequencing inferred from the sample’s viral load as indicated by the diagnosis Ct value. Such a strategy ultimately avoided NGS of some samples that may have been critical in reconstructing the patterns of spread of A(H1N1)pdm09 virus and persistence of transmission clusters.

In conclusion, although the intensity of influenza surveillance in Africa still lags behind that of other continents, our findings suggest that considerable influenza virus diversity circulates within the continent, including virus lineages that are unique to the region, as reported for Kenya; these lineages may be capable of dissemination to other continents through a globally migrating virus population. Further knowledge of the viral lineages that circulate within understudied tropical and subtropical regions is required to understand the full diversity and global ecology of influenza viruses in humans and to inform vaccination strategies within these regions.

## Data Availability

All generated sequence data were deposited in the Global Initiative on Sharing All Influenza Data (GISAID).

https://github.com/DCollinsOwuor/H1N1pdm09_Kenya_Phylodynamics/tree/main/Data/.

## SUPPORTING INFORMATION

### DISCLOSURE

The findings and conclusions in this report are those of the authors and do not necessarily represent the official position of the Centers for Disease Control and Prevention.

### FUNDING

The authors D.C.O. and C.N.A. were supported by the Initiative to Develop African Research Leaders (IDeAL) through the DELTAS Africa Initiative [DEL-15-003]. The DELTAS Africa Initiative is an independent funding scheme of the African Academy of Sciences (AAS)’s Alliance for Accelerating Excellence in Science in Africa (AESA) and supported by the New Partnership for Africa’s Development Planning and Coordinating Agency (NEPAD Agency) with funding from the Wellcome Trust [107769/Z/10/Z] and the UK government. The study was also part funded by a Wellcome Trust grant [1029745] and the USA CDC grant [GH002133]. N.F.M. is supported by the Swiss National Science Foundation (PZEZP3_191891). This paper is published with the permission of the Director of KEMRI.

## SUPPLEMENTARY FIGURE LEGENDS

**S1 Figure:**
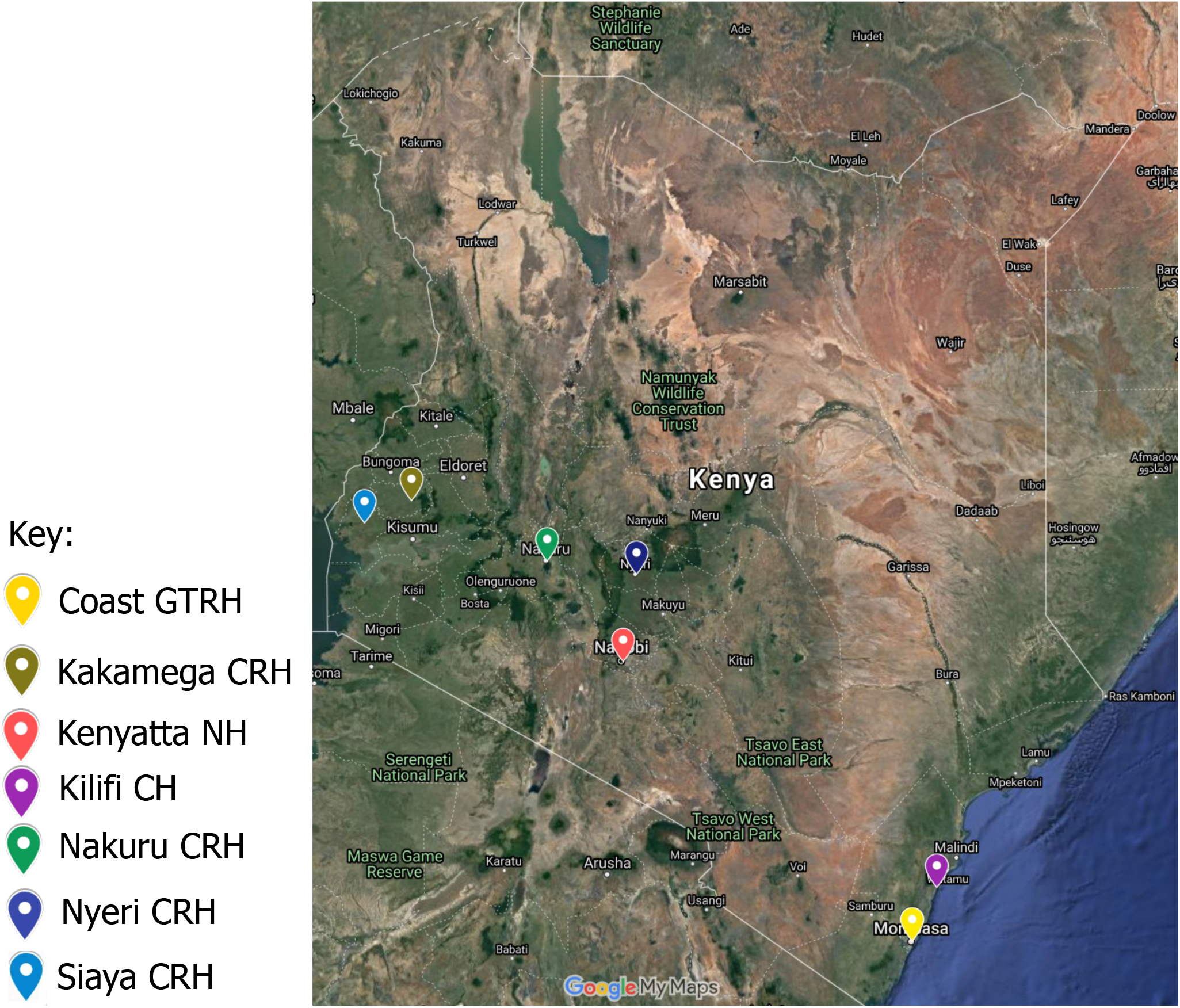
Map of Kenya showing the influenza sentinel surveillance sites for SARI used in this study. SARI, Severe Acute Respiratory Illness; GTRH, General Teaching and Referral Hospital; CH, County Hospital; CRH, County and Referral Hospital; NH, National Hospital.

**S2 Figure:**
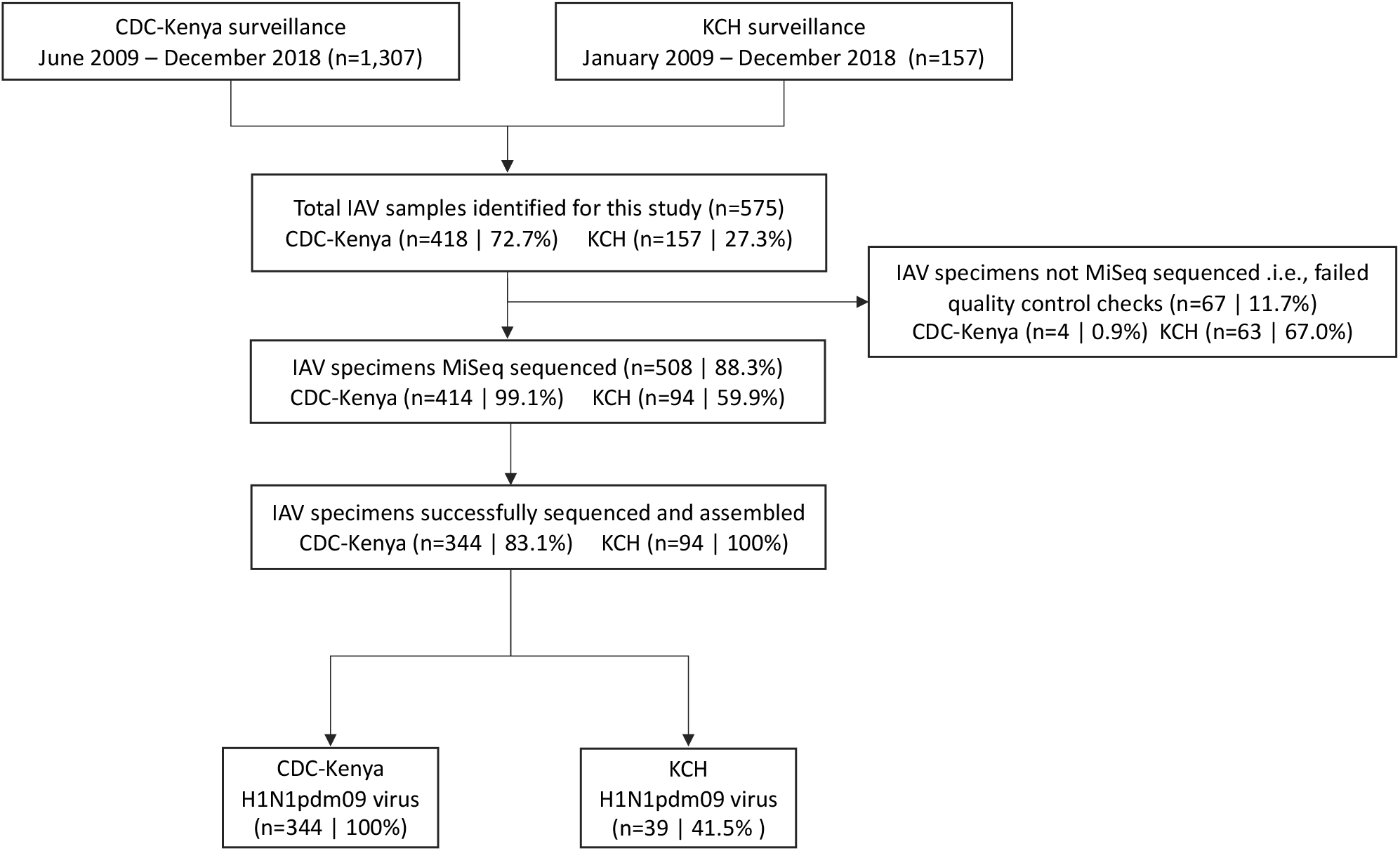
Sample processing flow for CDC-Kenya and KCH surveillance of IAV positive specimens, 2009-2018. Next-generation sequencing generated 344 and 39 codon-complete A(H1N1)pdm09 virus sequences from the CDC-Kenya and KCH surveillance studies, respectively, which were used for this report. CDC, Centers for Disease Control; KCH, Kilifi County Hospital; IAV, influenza A virus.

